# Associations of epigenetic aging with cortical thickness, brain age gaps, and neuroanatomical heterogeneity

**DOI:** 10.1101/2025.11.09.25339844

**Authors:** Marco Hermesdorf, Jan Homann, Jan Ernsting, Marisol Herrera-Rivero, Tim Hahn, Christina Lill, Klaus Berger

## Abstract

The biological aging process exhibits heterogeneous effects on different tissues, manifesting as tissue-specific variations in structural integrity and functional decline. Previously developed models are able to predict age from DNA methylation in the blood and the difference between estimated epigenetic age and chronological age is suggested to reflect accelerated or decelerated biological aging. While most prior studies have focused on the association between epigenetic age acceleration and global cortical thickness, it remains to be determined whether biological aging varies across specific cortical regions. This study aimed to assess associations between epigenetic age acceleration and regional cortical thickness, brain age gaps, as well as intra- and interindividual neuroanatomical heterogeneity in 756 participants of the BiDirect Study, including 430 participants from the general population and a cohort of 326 individuals with depression. Epigenetic age was estimated from whole blood DNA methylation data using the GrimAge algorithm. We observed an association of epigenetic age acceleration with cortical thinning across almost all cortical regions, suggesting a global association without regional confinement. This result was additionally underpinned by showing that accelerated epigenetic aging was also associated with increased interindividual neuroanatomical heterogeneity in contrast to a lack of association between epigenetic aging acceleration and intraindividual neuroanatomical heterogeneity. Accelerated epigenetic aging was furthermore paralleled by higher neuroimaging-based brain age gaps, suggesting at least partly shared aging processes. Together, these findings highlight that accelerated epigenetic aging reflects a global rather than region-specific neuroanatomical aging process, linking molecular and structural markers of brain aging and underscoring the potential of epigenetic clocks as biomarkers for brain health and neurodegenerative risk.

## Introduction

Aging is a universal process that affects almost all biological entities and is characterized by a progressive decline in the anatomical integrity of organs and their biochemical functioning^1^. The aging process and the continuous accumulation of detrimental physiological changes that it is composed of are associated with increased risks for the development of diseases as well as mortality, particularly with regard to neurodegenerative diseases such as Alzheimer’s, cardiovascular diseases, and cancer^2^. Importantly, chronological age as a non-modifiable risk factor and biological age are not the same as individuals can differ in the latter despite having the same chronological age. Biological aging is partly modifiable and can vary substantially based on genetics and lifestyle behaviors such as smoking, physical activity, and dietary patterns^3^. However, biological aging is a complex and multifaceted concept without a universally accepted consensus on the definition. It can be examined on a tissue-specific level, allowing for the monitoring of disease-associated processes affecting both individual tissues as well as the entire body.

One biological process that is highly sensitive to aging is DNA methylation, which is an epigenetic modification where a methyl group is added to cytosine-phosphate-guanine (CpG) sites of the DNA, a process that affects gene expression^4,5^. With advanced statistical models, the age-sensitive patterns of DNA methylation, commonly measured in blood, can be used to derive so-called “epigenetic clocks” as an estimate of biological age. The difference between chronological and biological age indicates accelerated or decelerated biological aging^6^. While the first generation of clocks were trained on chronological age, the GrimAge clock^7^ as a second-generation clock furthermore incorporates predictors of mortality, including smoking pack years and circulating proteins related to cardiovascular risk and inflammation, thereby better capturing functional decline and mortality compared to first-generation clocks^8^. The brain is also highly sensitive to aging patterns across the entire lifespan showing continuous changes in gray matter volume and cortical thickness^9^. Hence, biological age can also be estimated for the brain by using voxel- or vertex-wise data commonly derived from T1-weighted magnetic resonance imaging (MRI) to train machine learning or deep learning models that capture aging trajectories and individual deviations thereof, so-called brain age gaps, that indicate accelerated or decelerated brain aging^10^.

Only a few studies investigated associations between epigenetic age and regional or vertex-wise cortical thickness in the general population. Two studies based on small sample sizes (n = 82 and n = 79, respectively) reported an inverse relationship between advanced epigenetic age and cortical thinning that was accentuated in frontal and temporal lobe regions^11,12^. A further investigation in population-based cohorts assessed only mean cortical thickness across the cortex and found an inverse association between accelerated epigenetic aging and globally lower cortical thickness^13^. Furthermore, evidence on associations between brain aging and accelerated epigenetic aging is scarce. One study found that accelerated brain age was associated with accelerated epigenetic age^14^. In addition, investigating how epigenetic aging might relate to the variability in global neuroanatomy, which can be assessed between-subjects (i.e. interindividual) with a person-based similarity index^15^ (PBSI) and within-subjects (i.e. intraindividual) using a coefficient of variation ratio^16^ (CVR), can further contribute to the understanding of how systemic biological aging manifests in the brain at the individual level that extends beyond commonly used measures of brain structure. However, to the best of our knowledge, variability in global neuroanatomy has not yet been assessed in relation to epigenetic aging. Importantly, variations in cell composition can substantially affect DNA methylation patterns and consequently the accuracy of biological age estimation^17^. However, not all studies that previously evaluated associations with neuroimaging-derived phenotypes adjusted their analyses for immune cell composition.

In the present study, we comprehensively assessed associations between accelerated epigenetic aging and regional cortical thickness, interindividual as well as intraindividual neuroanatomical variability as well as brain age gaps. We hypothesized that advanced epigenetic age was related to lower cortical thickness across the brain as well as to accelerated brain aging and higher neuroanatomical variability.

## Methods

### Participants

The current study analyzed data from individuals recruited as part of the BiDirect Study conducted in Münster (Germany). Participants from the general population aged between 35 and 65 years were invited to take part in the baseline examination program following the selection of a random sample from the local population register. In addition, individuals in the same age range with a diagnosed current or recurrent episode of major depression (ICD-10 F32 or F33) under in- or outpatient treatment were also invited to participate in the examination program. In the course of the BiDirect examination program, participants underwent an extensive examination program including a face-to-face interview covering lifestyle and previous physician-diagnosed diseases, and a physical examination program. Participants also received MRI, filled out self-report questionnaires, and blood samples were drawn. The population-based cohort comprised a total of 911 participants and the depression cohort included 984 patients. Population-based controls (n = 239) and patients with depression (n = 268) without available 3D-T1 MRI were excluded as were population-based controls (n = 23) and patients with depression (n = 21) whose MRI scans did not pass quality control after running an in-house script and visual inspection. Those from the population-based cohort (n = 14) as well as patients with depression (n = 17) without available brain age data were excluded. Controls (n = 205) and patients with depression (n = 352) without available epigenetic data were also removed from the sample. The final sample consisted of 430 population-based controls and 326 patients with depression. All participants provided written informed consent, the BiDirect Study was conducted in compliance with the Declaration of Helsinki and approved by the local ethics committee.

### Magnetic resonance imaging

Structural 3D-T1-weighted turbo field echo images were obtained with the following specifications: TR = 7.26 ms, TE = 3.56 ms, flip angle = 98°, matrix size = 256 x 256, FOV = 256 x 256 mm², 160 sagittal slices, a slice thickness of 2 mm (scanner-wise reconstructed to 1 mm), resulting in a final voxel size of 1 x 1 x 1 mm³.

The data were processed using SPM12 (revision 7771; fil.ion.ucl.ac.uk/spm/software/spm12/) and CAT12.8.226^18^ (neuro-jena.github.io/cat/) toolboxes running on Matlab R2023a (MathWorks, Natick, MA, USA). The images underwent bias correction and were segmented into gray and white matter, and cerebrospinal fluid. Following affine registration to the Montreal Neurological Institute (MNI) standard space, the imaging data were processed through a multi-step segmentation pipeline that included local adaptive segmentation, adaptive maximum a posteriori segmentation, and partial volume estimation. Gray matter volumes for the caudate nucleus, putamen, thalamus, nucleus accumbens, pallidum, amygdala, and the hippocampus were extracted according to the Neuromorphometrics atlas. The images were then brain-extracted and non-linearly registered to MNI space. CAT12 uses a projection-based approach^19^ for the estimation of cortical thickness as the distance between the gray-white matter boundary and the pial surface. Mean cortical thickness estimates for 68 regions were derived based on the Desikan-Killiany atlas.

### Interindividual neuroanatomical variability: Person-based similarity index

Between-subject neuroanatomical variability was operationalized as PBSI^15^, which was derived by first computing Pearson correlations of the concatenated regional cortical thickness and gray matter volume data for each pair of participants separately for each cohort, yielding *n* - 1 correlation coefficients per individual which were subsequently averaged to obtain one single index score per person. The PBSI scores reflect the degree of neuroanatomical variability whereas higher scores indicate greater similarity or homogeneity, and lower scores reflect greater between-subject or interindividual neuroanatomical heterogeneity with reference to all other individuals.

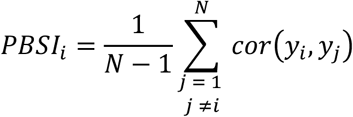

### Intraindividual neuroanatomical variability: Coefficient of variation

The CVR was used as an estimate of within-subject or intraindividual neuroanatomical variability. The CVR was calculated per individual as the standard deviation across their own atlas-based regional cortical thickness estimates and divided by the mean of these, expressed in percentage points. A higher CVR indicates greater intra-individual neuroanatomical heterogeneity.

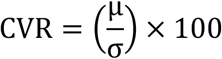

### Brain age gaps

The brain age gaps were predicted using the publicly available MCCQRNN brain age model^10^, trained on 30,115 individuals from the German National Cohort (NAKO). The sample consisted of 16,830 male and 13,285 female subjects with median age of 49 years (min: 19, max: 74, mean: 48.24, std: 12.28). The training procedure was identical to the baseline MCCQRNN model: After MRI preprocessing, the images were resampled into 3-mm isomorphic voxels, and gray matter voxels were extracted and vectorized using the PHOTON AI software. Finally, the MCCQRNN model was applied. The resulting predictions account for two sources of uncertainty: aleatory and epistemic uncertainty. Aleatory uncertainty is contained in the training data caused by variance or noise during measurement. Epistemic uncertainty is caused by the model or absence of training samples in out of distribution settings. The model predicts 101 quantiles per sample and combines aleatory and epistemic uncertainty in these quantiles. For subsequent analysis, the median value of the predicted quantiles per subject is used. Brain age gap is then calculated by subtracting the described prediction from the chronological age.

### DNA methylation

DNA methylation (DNAm) profiling was performed on the “Infinium MethylationEPIC” v1.0 or v2.0 array (Illumina, Inc.) on aliquots of DNA extracts at ∼50ng/µl concentration. Data pre-processing was done with R 4.1.2 and the R-package “Bigmelon” using recommended default parameters for the quality control (Gorrie-Stone et al., 2019). To this end, the samples from the first and second batch were loaded and pre-processed separately. Briefly, probes were filtered according to the detection P value. Probes with more than 1% of samples having a detection P value of 0.05 were removed from the analysis, as well as probes with a bead count smaller than 3 in more than 5% of the samples. Then, outliers were identified with the *outlyx* function based on two times the interquartile range of the first PC of the DNA methylation data and the *pcout* function with a threshold of 0.15. *Pcout* is a function to identify outliers computationally fast in high-dimensional data (Filzmoser et al., 2008). Additionally, samples with a bisulfite conversion efficiency below 80% as estimated by the *bscon* function were removed. The samples were reloaded with outliers excluded and normalized with the function *dasen*. The function *qual* was used to determine the extent of change in beta values in each sample due to normalization. A very large change is indicative of an outlying sample and should therefore be excluded from the analysis. Samples with a root-mean-square deviation of 0.1 or larger were removed and loading and normalization were repeated with the new sample set with removed outliers. After quality control and filtering, X samples remained, for which X had age and sex information required for the GrimAge calculation available. GrimAge calculation was performed using the “DNA Methylation Age Calculator” (https://dnamage.genetics.ucla.edu/new) according to the manual from the website: ManualEpigeneticClock3_new.pdf. As described in the manual, non-filtered, raw DNAm data were used, and the tissue annotation was set to “Blood WB”. Cell-type estimations were made for granulocytes (Gran), monocytes (Mono), B cells (Bcell), CD4+ T cells (CD4T), CD8+ T cells (CD8T) and natural killer (NK) cells according to the method by Houseman^20^.

### Statistical analysis

A GrimAge age gap estimate (GAGE) indicating accelerated or decelerated epigenetic aging was operationalized as the residuals derived from a model where the estimated epigenetic age from the DNA methylation data was regressed on chronological age. The GAGE was used as independent variable for the analyses of regional cortical thickness data with regression models. The associations between brain age gaps, PBSI, and CVR scores as respective dependent variables and GAGE as independent variable were also analyzed with separate multiple regression models. Brain age gaps were calculated as the difference between estimated brain age (mean aleatory epistemic) and chronological age. All analyses were adjusted for age, sex, cohort, DNA methylation batch, and the following cell type estimates: CD8 T cells, CD4 T cells, natural killer cells, B lymphocytes, monocytes, and granulocytes. The analyses of regional cortical thickness and gray matter volume data were corrected for multiple comparisons by using the Benjamini-Hochberg false discovery rate procedure^21^. Statistical analyses were conducted with R (version 4.31).

## Results

Sample characteristics are shown in Table 1. A higher GAGE was significantly associated with lower cortical thickness bilaterally across all lobes of the brain as shown in Table 2 and Figure 1. Effect sizes in terms of variance explained for cortical thickness were most pronounced in the temporal and parietal lobes (Figure 1). Beta estimates as shown in Figure 2 indicated the strongest associations of higher GAGE with lower cortical thickness in the caudal and rostral parts of the right anterior cingulate cortex. Regarding gray matter volume, a higher GAGE was exclusively related to a lower volume of the thalamus and the globus pallidus (Table 2), bilaterally. Furthermore, individuals with a higher GAGE also exhibited a significantly higher brain age gap (*β* = 0.11, 95% confidence interval 0.039 to 0.175, *p* = 0.002, partial η^2^ = 0.01). A partial residual plot showing the adjusted association between GAGE and brain age gaps is shown in Figure 3. GAGE was weakly but significantly associated with PBSI, showing that accelerated epigenetic aging (*β* = - 0.0003, 95% confidence interval -0.0005 to -0.0001, *p* = 0.001, partial η^2^ = 0.01) was parallelled by a lower PBSI. The p-value was obtained using 1000 bootstrap iterations due to a ceiling effect of PBSI scores. We did not observe significant associations between GAGE (*p* = 0.64) and CVR as a measure of intraindividual neuroanatomical heterogeneity.

**Figure 1.**
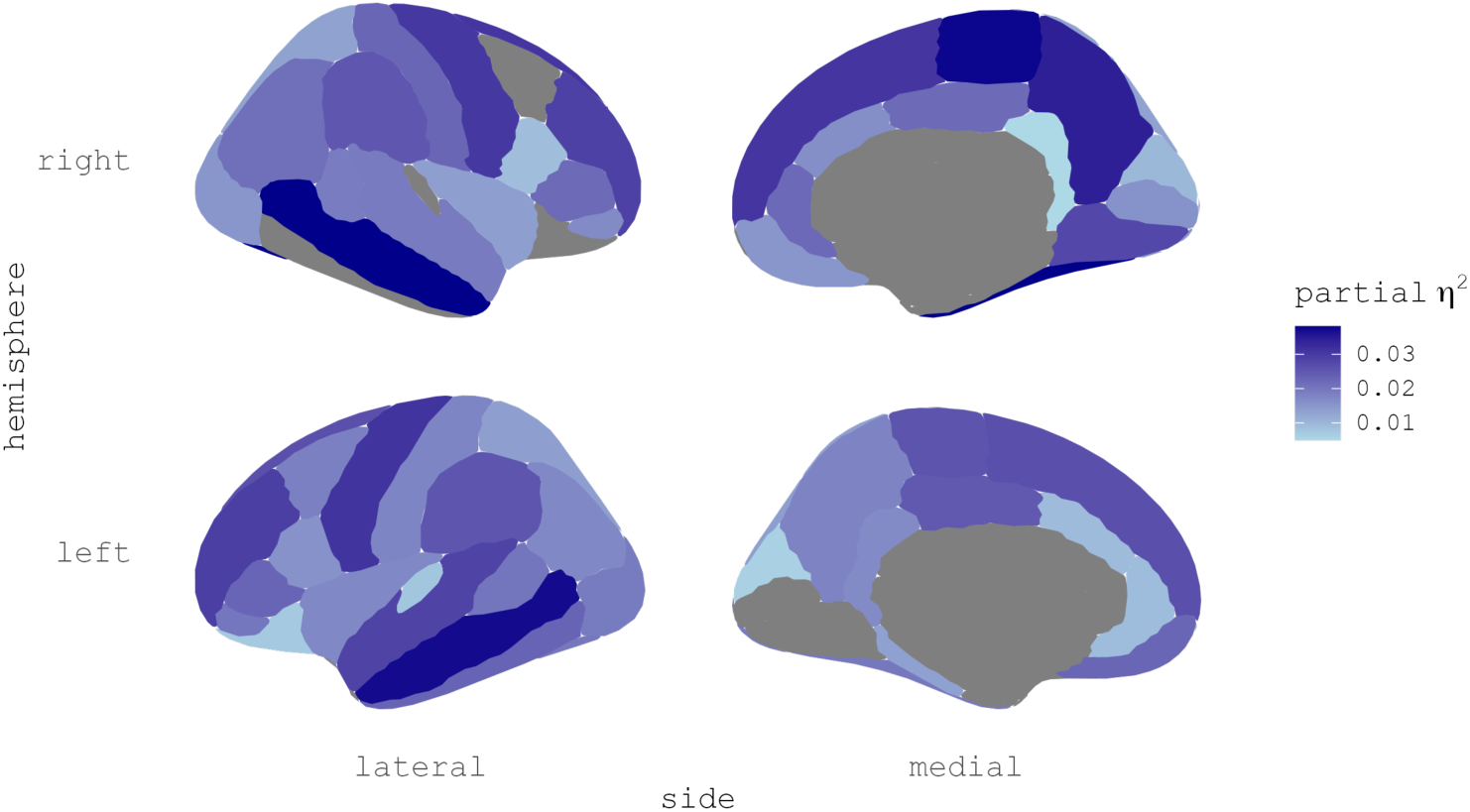
Associations of accelerated epigenetic aging with lower cortical thickness. Explained variance for cortical regions significantly associated with the GrimAge age gap estimate. Effect sizes are presented as partial η^2^.

**Figure 2.**
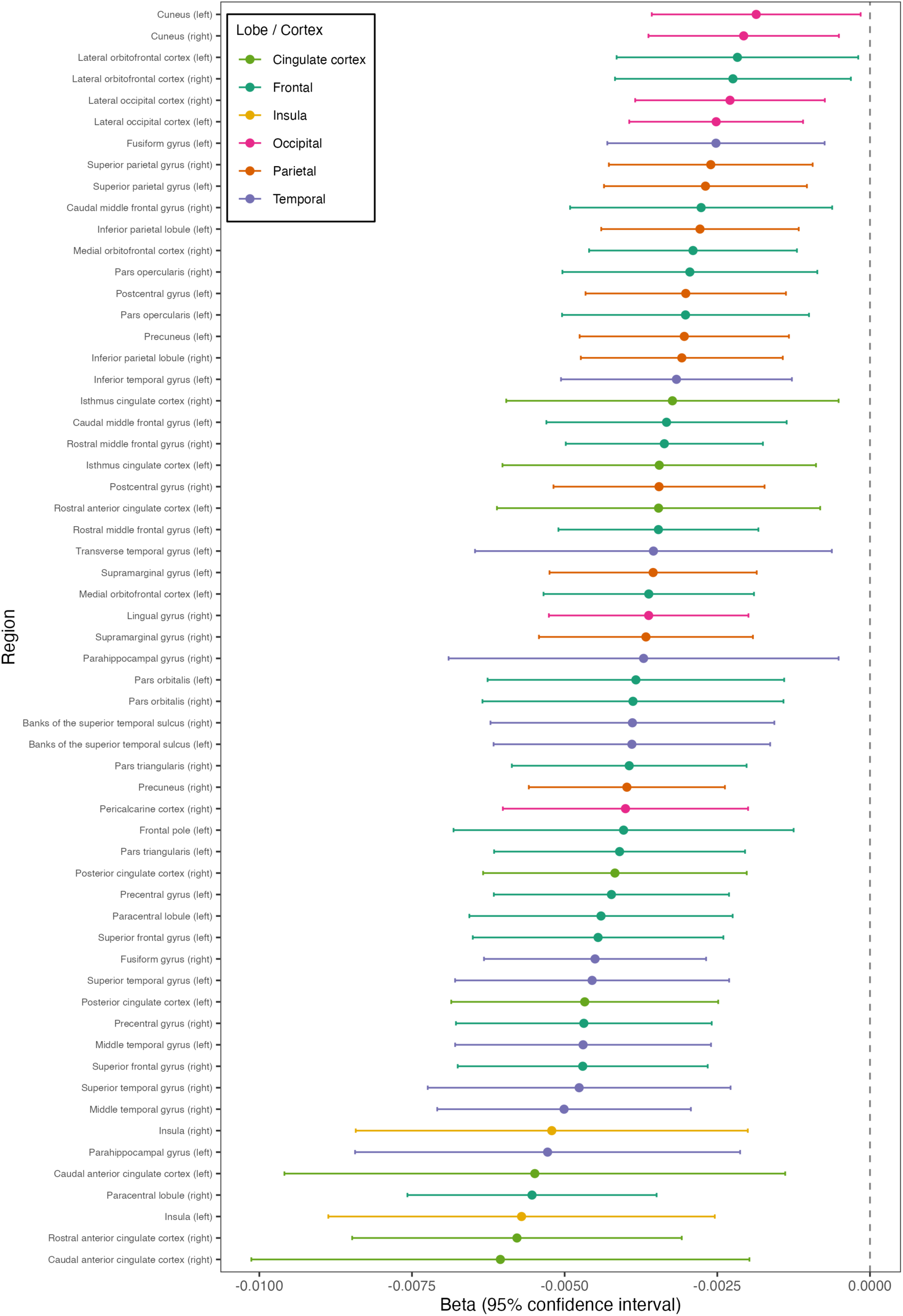
Beta estimates and confidence intervals for the GrimAge age gap estimates for regions significantly associated with cortical thickness.

**Figure 3.**
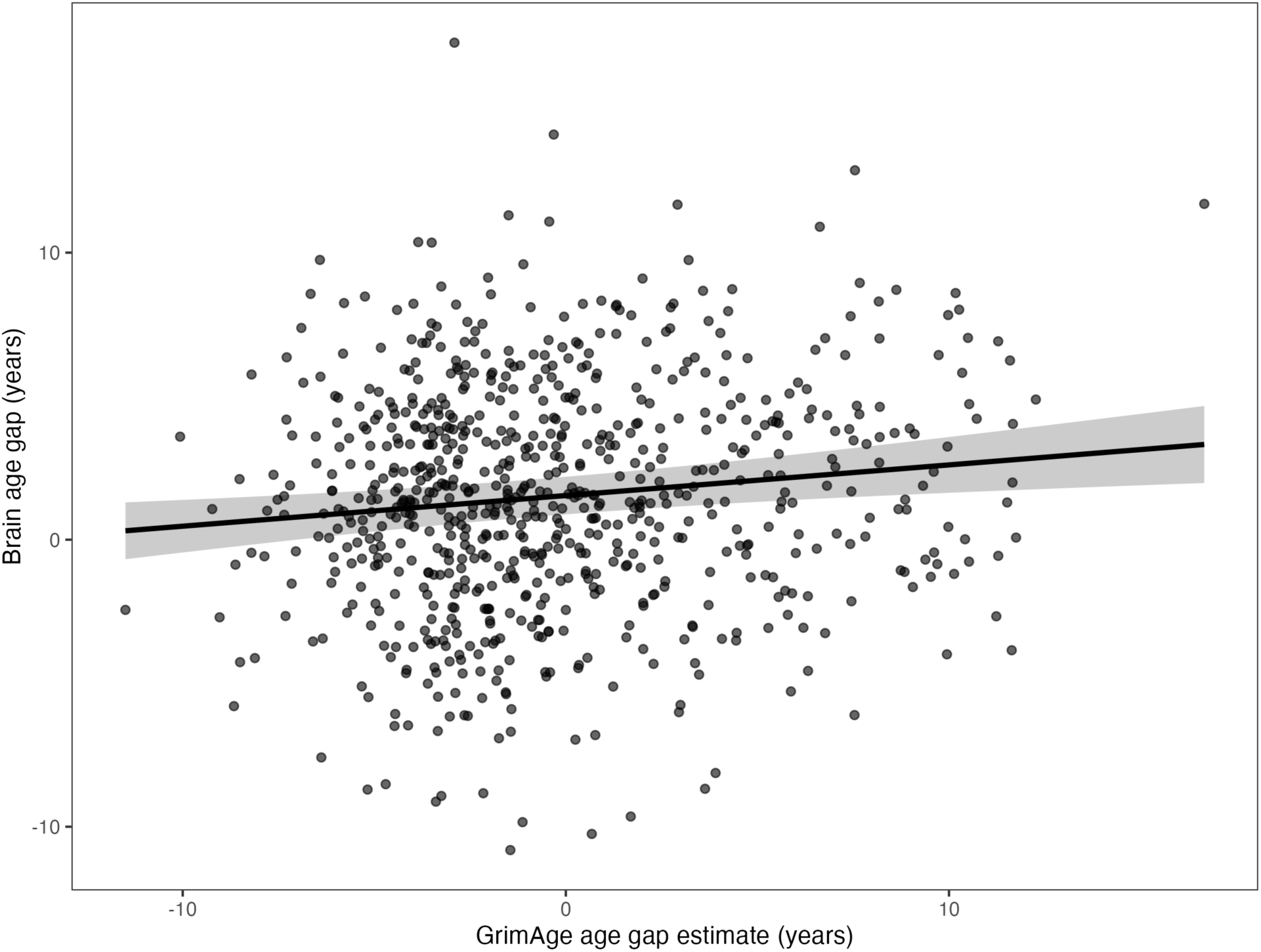
Partial residual plot of the association between the GrimAge age gap and brain age gap.

**Table 1.** Sample characteristics.

|  | General population<br>(N = 430) | Depression<br>(N = 326) |
| --- | --- | --- |
| Sex |  |  |
| Female | 229 (53.3%) | 191 (58.6%) |
| Male | 201 (46.7%) | 135 (41.4%) |
| Age (years) |  |  |
| Mean (SD) | 53.2 (7.81) | 50.5 (7.19) |
| Median [Min, Max] | 54.2 [35.5, 66.1] | 50.7 [35.4, 65.5] |
| Epigenetic age (years) |  |  |
| Mean (SD) | 66.3 (13.1) | 62.5 (12.0) |
| Median [Min, Max] | 66.3 [39.5, 93.8] | 60.3 [38.4, 91.8] |

**Table 2.**
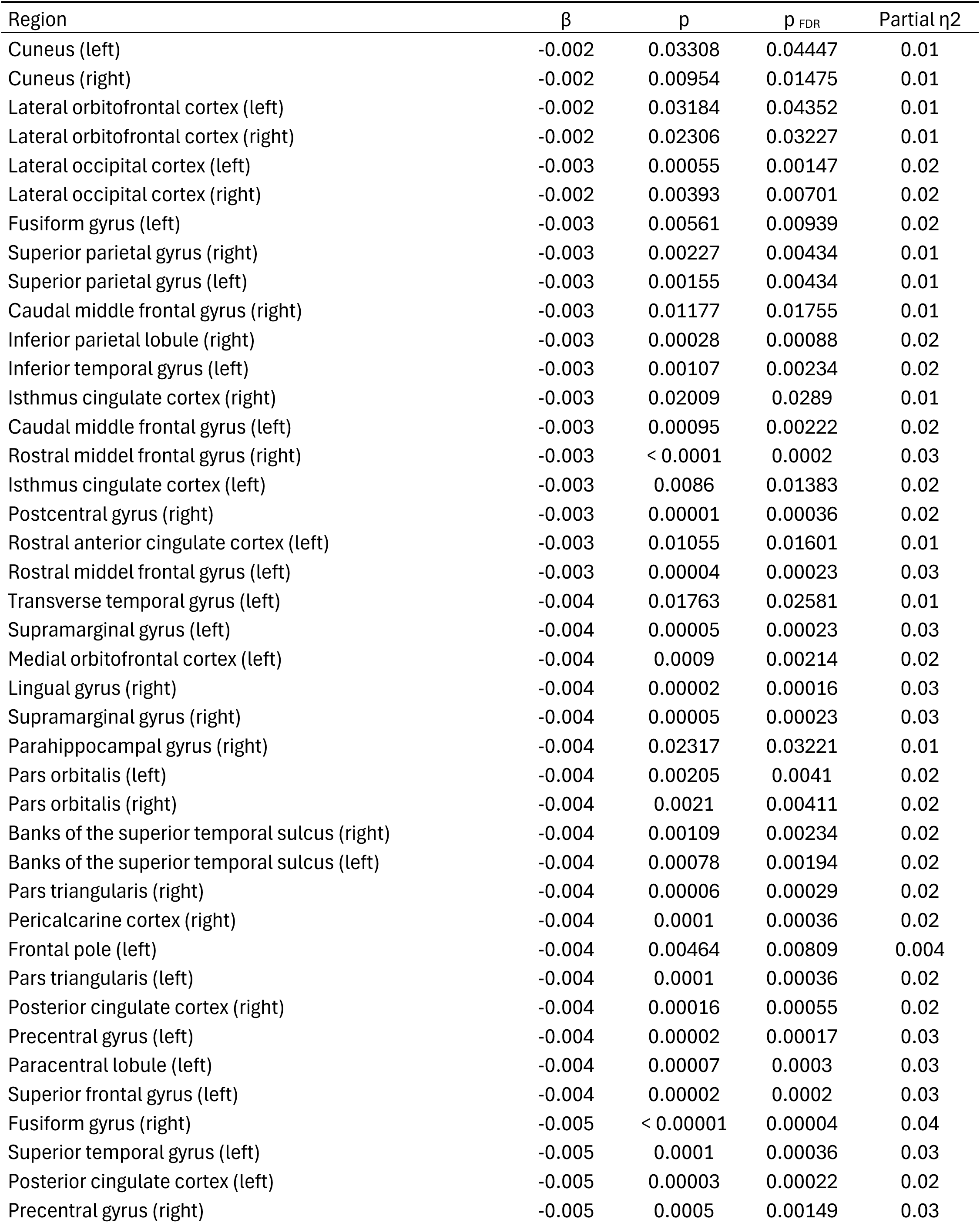
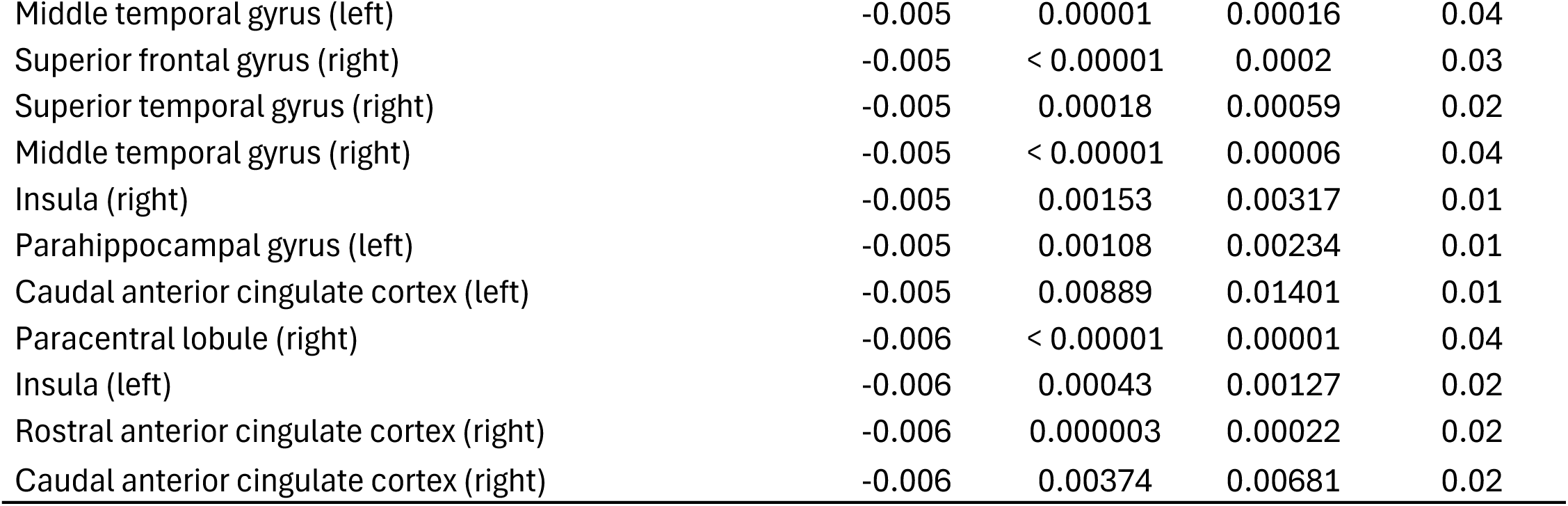

## Discussion

This study analyzed the association between epigenetic aging and cortical thickness, brain age gaps, and neuroanatomical heterogeneity. Our results demonstrate that accelerated epigenetic aging is significantly associated with lower cortical thickness in all lobes of the brain. We also observed that participants with an accelerated epigenetic age had an accelerated brain age of 0.11 years per year of accelerated epigenetic age. In addition, we found that accelerated epigenetic aging was related to higher interindividual but not intraindividual neuroanatomical heterogeneity.

Previously, it has been shown that accelerated epigenetic aging, also assessed with the GrimAge clock, was associated with lower mean cortical thickness across the brain^13^. The current results add that accelerated epigenetic aging estimated from blood samples is consistently related to widespread cortical thinning in the brain in almost all cortical regions. The finding of a global effect without any particular regional confinement is further supported by the interindividual neuroanatomical heterogeneity that was associated with accelerated epigenetic aging in contrast to the intraindividual neuroanatomical heterogeneity that was not related to epigenetic aging. The GrimAge clock as a second-generation clock incorporates predictors of mortality^7,8^ in contrast to the first-generation clocks, which were trained to merely predict chronological age. A prior study using a consensus model of first-generation clocks found only small clusters in the brain where accelerated epigenetic aging was associated with cortical thinning, although without correcting for multiple comparisons^11^. Another study^12^ using a first-generation clock reported more widespread cortical thinning but did not control for chronological age in their analysis of epigenetic age acceleration, which could introduce bias towards the null hypothesis^22^. Our findings of a higher brain age gap associated with accelerated epigenetic aging are consistent with a previous study^14^ that employed the first-generation Horvath clock^6^ using saliva samples. The current study thus shows that accelerated epigenetic aging estimated from the blood as a systemic tissue corresponds to accelerated brain aging that was assessed using MRI.

Previously, studies have shown that DNA methylation is potentially reversible by the introduction of specific encoding transcription factors^23,24^. Epigenetic aging is also modifiable through lifestyle interventions^25,26^ targeting diet, physical activity, and sleep which could potentially delay the onset of age-related diseases or functional decline in late life. Since DNA methylation affects gene expression, the accumulating pattern of DNA methylation characteristic of accelerated epigenetic aging could hence constitute one of many mechanisms of how lifestyle behavior ultimately affects disease-related outcomes.

The present study comes with a few limitations. The sample includes participants aged between 35- and 65-years during recruitment, which might limit generalizability to older adults where aging-related mortality and functional decline steadily increase. While it has been shown that age-related thinning of the cortex is linear even beyond the age of 65 years^9^, this does not exclude the possibility that cascading accelerated epigenetic aging in late-life could manifest in compounded patterns of cortical thinning. Furthermore, we estimated epigenetic age in DNA isolated from EDTA blood and not directly in neural tissue, which could potentially underestimate the relationship between DNA methylation and cortical thickness. However, while sampling neural tissue to estimate epigenetic age would be unfeasible to conduct in the general population, the present results suggest that blood-based DNA methylation is a pragmatic and expedient systemic marker indicative of brain alterations related to accelerated aging.

In conclusion, the present study demonstrated that accelerated epigenetic aging as measured in the blood is a viable indicator of interindividual neuroanatomical heterogeneity, accelerated brain aging, and cortical thinning across the brain without any particular regional confinement. While different tissues clearly exhibit distinct aging phenotypes, there remains a shared and fundamental process that underpins its various manifestations throughout the body.

## Data Availability

The data analyzed in the present study are available upon reasonable request.

## Conflicts of interests

The authors have no conflicts of interest to declare.

## Authorship contributions

Conception of this analysis: M.H. Conception and design of the BiDirect Study as data source: K.B. Obtained funding: K.B. Estimation of epigenetic age: J.H., C.L. Estimation of brain age: J.E., T.H. Processing of cortical thickness data and analysis: M.H. Drafting the manuscript: M.H. Critical revision of the manuscript: K.B., M.H.-R., C.L., J.H., J.E., T.H.

## Acknowledgements

The current work was supported by grants from the German Federal Ministry of Education and Research (BMBF; grants FKZ01ER0816 and FKZ-01ER1506) to K.B. and J.E. was supported by the Medical Scientist Kolleg InFlame funded by the Else Körner-Fresenius Foundation.

## Notes

### Competing Interest Statement

The authors have declared no competing interest.

### Author Declarations

All participants provided written informed consent, the BiDirect Study was conducted in compliance with the Declaration of Helsinki and approved by the local ethics committee of the University Hospital Muenster as well as the ethics committee of the medical chamber of the Westphalia-Lippe (Germany).

